## Supplemental Appendix for "Proposed Context-of-Use Evaluation Framework for Medication Management Tasks Completed by Generative Artificial Intelligence"

Interrater reliability - Krippendorff's Alpha

PNA CMM interrater agreement (Krippendorff's Alpha)
============================================================
Cases aligned across 3 annotators: 150
Bootstrap iterations: 2000

Identification of Relevant Information: alpha=0.217 [0.112, 0.318]
Therapeutic Goal: alpha=0.208 [0.131, 0.289]
Medication Therapy Problem: alpha=1.000 [1.000, 1.000]
Medication Therapy Regimen Accuracy: alpha=0.601 [0.484, 0.702]
Clinician Preference: alpha=0.577 [0.469, 0.662]
Medication Therapy Format: alpha=1.000 [1.000, 1.000]
Monitoring Parameters: alpha=-0.207 [-0.258, -0.158]
Duration: alpha=0.435 [0.221, 0.607]
Automatic Failure - Serious or Severe Harm (put a 1 here if meeting failure criteria, put a 0 if no failure): alpha=0.027 [-0.079, 0.146]
Automatic Failure - Hallucinated Information (put a 1 here if meeting failure criteria, put a 0 if no failure): alpha=0.152 [0.042, 0.252]
